# Adverse outcomes and associations of dengue in pregnancy – a retrospective cohort study

**DOI:** 10.1101/2024.04.07.24304931

**Authors:** Oshadhi Lathmini Nallaperuma, Hematha Malinath Senanayake, Janaka Chandana Godevithana

**Author notes:** Postal address -Geethani, Udukawa, Denipitiya, Sri Lanka. Lecturer, Department of Obstetrics and Gynaecology, Faculty of Medicine, Sabaragamuwa University of Sri Lanka, Belihuloya, Sri Lanka (Oshadhi Lathmini Nallaperuma).

## Abstract

**Objective:** Dengue infection is a significant health issue in tropical areas over the globe. Physiological changes of pregnancy can be confounding the clinical presentation of dengue in pregnancy. This study aimed to investigate the adverse outcomes of antenatal dengue infection and factors associated with maternal, fetal, and neonatal complications.

**Methods:** A total of 115 pregnant women with laboratory-confirmed dengue infection admitted to two tertiary referral hospitals in Sri Lanka were followed from admission until delivery and discharge. Data on clinical and laboratory outcome variables were collected and analyzed using descriptive and inferential statistics.

**Results:** Dengue fever accounted for the majority of cases (63.5%), followed by dengue hemorrhagic fever (33.9%) and, dengue expanded syndrome (2.6%). Maternal adverse outcomes were observed in 27.8% of cases with 19.1% of ICU admissions. Dengue category, trimester of infection, platelet count, and AST levels showed a significant association with adverse maternal outcomes. Fetal adverse outcomes affected 8.1% of cases, while neonatal adverse outcomes occurred in 28.7% of live births, with transplacental transmission observed in two cases.

**Conclusion:** Dengue infection during pregnancy can lead to significant maternal, fetal, and neonatal morbidity and mortality. Identifying risk factors may help predict the severity of these outcomes. Further research is needed to develop robust prediction models and improve management strategies.

## Introduction

The global burden of dengue infections presents a significant public health challenge, with an estimated 100 to 400 million cases occurring annually worldwide, endangering approximately half of the world’s population.^1^ Sri Lanka stands as a dengue-endemic country, grappling with an average caseload ranging from 50,000 to 150,000 cases per year.^2^ Notably, in 2017, dengue emerged as the leading cause of maternal mortality in Sri Lanka, accentuating the critical importance of understanding and addressing the impact of this vector-borne viral infection during pregnancy.^3^

Dengue virus has four distinct strains named DEN1 to DEN4. The primary vector responsible for dengue transmission is the *Aedes aegypti* mosquito.^4^ Despite the existence of comprehensive guidelines from the World Health Organization (WHO) and local health authorities on the management of dengue, the disease remains a formidable challenge due to its varied clinical manifestations and potential complications.^1,5^

Classification of dengue encompasses three distinct clinical entities: dengue fever (DF), dengue hemorrhagic fever (DHF) with or without shock, and dengue expanded syndrome (DES). Furthermore, the disease progresses through distinct phases, including the febrile phase, critical phase, and recovery phase. In some cases, complications may arise, leading to dengue shock syndrome (DSS), which significantly increases mortality and morbidity rates.^6^

Pregnancy accentuates the complexities of managing dengue due to the distinctive physiological changes occurring. Peripheral resistance drops by 40%, while plasma volume increases by 50-60% in the late third trimester. Extracellular volume experiences a 16% rise, accompanied by reductions in packed cell volume and platelet counts, and an increase in white cell count during the peri-partum period.^7^ These alterations complicate the diagnosis and management of dengue in pregnancy, given that key features of dengue infection include thrombocytopenia and extracellular fluid loss.^1^

Management of dengue primarily relies on literature focused on non-pregnant adults and expert opinions. This study aims to investigate the adverse maternal, fetal, and neonatal outcomes of dengue infection during pregnancy and any factors that may be associated with these adverse outcomes.

## Materials and Methods

This is a retrospective cohort study without a matched comparator group. The cohort of pregnant women with laboratory-confirmed dengue infection, identified either by a positive non-structural protein 1 (NS1) antigen test or dengue-specific immunoglobulin M and G (IgM and IgG) tests, between January 1, 2017, and December 31, 2017, who were admitted to De Soysa Hospital for Women (DSHW) and the National Hospital of Sri Lanka (NHSL), were included in the study. DSHW is a leading tertiary hospital for women receiving referrals from across the country. To minimize confounding bias, ectopic pregnancies, multiple pregnancies, and pregnancies with pre-identified obstetric or fetal complications were excluded. Data were collected retrospectively using medical records and entered into a preprinted form. These cases were followed until the end of the pregnancy and discharge.

Data on characteristic symptoms, number of days on admission, highest recorded temperature during the episode, trimester, and category of dengue (DF, DHF, DES), lowest platelet count, rise of aspartate aminotransferase (AST) > 30IU/L, highest AST level, rise of alanine aminotransferase (ALT) >32IU/L, and the highest ALT level were collected. Characteristic symptoms considered in the study were; fever, arthralgia, myalgia, respiratory symptoms, retroorbital pain, headache, back pain, nausea and vomiting. Fever was defined as skin (axillary) temperature > 37.2 ^0^C.^8^ Trimesters were defined as follows; first trimester (T1) first day of last menstrual period to 13 weeks and 6 days, second trimester (T2) 14 weeks and 0 days to 27 weeks and 6 days, third trimester (T3) 28 weeks and 0 days to delivery.^9^ Pregnancy specific cut offs were utilized for laboratory tests.^10^ Outcomes of the acute infection and delivery outcomes both were collected and they were categorized as maternal, fetal and neonatal adverse outcomes. Possible confounders age and parity were also collected. Age was categorized as <20, 20-30, 31-40, >40, and parity as 0, 1-3, >3.

Statistical analysis was performed using the SPSS software version 22. Descriptive parameters were presented as mean, median, range and, percentage. Associations between categorical variables were assessed by Chi-squared test. However, all continuous variables did not follow a normal distribution. Therefore, an independent sample Mann-Whitney U test was used to determine the significance of the difference in medians between two groups: those with adverse outcomes and those without adverse outcomes. A P value of <0.05 was considered statistically significant.

There are no conflict of interests and this study has been self-funded by the researchers. The study has received approval from the Ethics Review Committee of the Faculty of Medicine, University of Colombo, Sri Lanka (EC/18/024).

## Results

During the study period, a total of 326 fever-related admissions were reviewed, from which 115 cases meeting the criteria were selected. Of these, 110 (95.7%) were successfully followed up until delivery, resulting in a loss to follow-up rate of 4.3%. The cases were distributed across the year, with 86 (74.8%) cases occurring between April and July. Among these cases, there were 73 (63.5%) instances of DF, 39 (33.9%) cases of DHF, and three (2.6%) cases of DES.

The commonest symptom was fever which was present in 110 (95.7%) patients. Myalgia and headache were the next, being present in 56 (48.7%) and 48 (41.7%) cases respectively. The recorded highest axillary temperature ranged from 37.2 ^0^C to 39.8 ^0^C. The fever had lasted an average of 4.1 days (median 4, range 1-7). Usually, an admission has occurred on day two of fever and the average hospital stay was 5.8 days (median 5, range 2-16). In the DHF category fluid leak had begun on an average of 4.2 days (median 4, range 2-6) after the onset of fever. Platelet counts dropped below 150,000/uL in 59 (80.8%) cases of DF and all cases of DHF, beginning to decline within an average of 2.5 days from fever onset. The lowest recorded platelet counts were 4000/uL in DF and 1000/uL in DHF. Transaminase levels rose in 48 (65.7%) cases of DF and 34 (87.1%) cases of DHF, starting to increase within an average of 3.3 days from fever onset (median 3, range 2-5).

### Adverse outcomes

Adverse outcomes were categorized as maternal, fetal and neonatal adverse outcomes [Table 01].

**Table 01.** Summary of adverse outcomes.

| Adverse outcome | DF | DHF | DES | Total | % <sup>†</sup> |
| --- | --- | --- | --- | --- | --- |
| **Maternal** |  |  |  |  |  |
| Maternal death | 0 | 1 | 0 | 1 | 0.86 |
| Intensive care unit admissions | 0 | 19 | 3 | 22 | 19.1 |
| Antepartum hemorrhage | 1 | 0 | 0 | 1 | 0.86 |
| Postpartum hemorrhage | 0 | 3 | 1 | 4 | 3.4 |
| Blood transfusion | 1 | 7 | 1 | 9 | 7.8 |
| Blood product transfusion | 0 | 5 | 1 | 6 | 5.9 |
| Precipitation of labour | 7 | 10 | 2 | 19 | 16.5 |
| Other | 0 | 3 | 3 | 6 | 5.2 |
| **Fetal** |  |  |  |  |  |
| First trimester miscarriages | 1 | 1 | 0 | 2 | 1.8 |
| Second trimester miscarriages | 0 | 2 | 0 | 2 | 1.8 |
| Intra uterine deaths | 1 | 2 | 2 | 5 | 4.5 |
| **Neonatal** |  |  |  |  |  |
| Meconium stained liquor | 5 | 4 | 0 | 9 | 8.9 |
| Neonatal intensive care unit admissions | 8 | 6 | 0 | 14 | 13.8 |
| Neonatal Dengue | 0 | 2 | 0 | 2 | 1.9 |
| Neonatal death | 0 | 0 | 0 | 0 | 0 |
| Low birth weight | 10 | 3 | 0 | 13 | 12.8 |
| Live births | 67 | 34 | 1 | 101 | 100 |

<sup>†</sup>The total number of cases in maternal, fetal and neonatal category were, 115, 110 & 101 respectively.
(DF – dengue fever, DHF – dengue hemorrhagic fever, DES- dengue expanded syndrome)

### Maternal adverse outcomes

Maternal adverse outcomes were observed in 32 (27.8%) cases, with 22 (19.1%) intensive care unit admissions. Serious adverse maternal outcomes included one maternal death, two cases of dengue hepatitis, and one case of dengue encephalitis, with the latter three attributed to DES.

Maternal death occurred due to delayed presentation with DSS in a 27-year-old female at a gestation of 19 weeks and five days. A history of fever was elicited. Dengue IgM and IgG tests were positive. A platelet count of 8000/uL and elevated transaminases (AST 11840 IU/L, ALT 1850 IU/L) were noted. patient experienced cardiorespiratory arrest on admission, leading to death on the same day after miscarrying the baby.

A case of dengue encephalitis presented with generalized tonic-clonic convulsions and tested positive for NS1 antigen, dengue IgM, and IgG in blood and cerebrospinal fluid. Following a slow recovery over two weeks, the patient had an uneventful antenatal period and delivered a healthy baby. The two cases of dengue hepatitis developed fulminant hepatic failure, with one progressing to hepatic encephalopathy.

Intensive care unit (ICU) admission was primarily due to the development of DHF. Among the three cases of post-partum hemorrhage (PPH) in the DHF category, one experienced massive blood loss necessitating insertion of a balloon tamponade and transfusion of six units of red cell concentrates and other blood products, with a platelet count of 39,000/uL during delivery. Other maternal complications in the DHF category included intrapartum maternal bradycardia, ventricular bigeminy, and bleeding manifestations such as hemoptysis and hematuria.

### Fetal adverse outcomes

The risk of developing a fetal adverse outcomes, excluding cases lost to follow-up, was 8.1% (9/110). Of the five cases of intrauterine fetal deaths, two were associated with dengue hepatitis, two with DHF, and one with DF. Additionally, two second-trimester miscarriages occurred, one coinciding with maternal death and the other with DHF at 14 weeks’ gestation.

### Neonatal adverse outcomes

There were 29 cases with neonatal adverse outcomes for 101 live births (28.7%). Reasons for neonatal intensive care unit admissions were for respiratory support in 10 cases, neonatal dengue in two cases, mother being in ICU in one case. The risk of transplacental transmission of dengue was 13.3% (2/15) when a live birth occurred during the dengue episode. One neonatal dengue infection occurred when the mother developed DHF at 40 weeks’ gestation, and the baby was delivered while the mother was still febrile, with the neonate exhibiting thrombocytopenia and testing positive for NS1 antigen, IgM, and IgG, but recovering without sequelae. The other neonatal dengue infection occurred when the mother developed DHF at 37 weeks and 3 days’ gestation, with the baby born at 38 weeks testing positive for NS1 antigen on the third day but not developing overt dengue infection, and subsequently being discharged healthy.

### Associations

Distribution of confounders were similar in both groups [Table 2]. Among categorical variables, (1) category of dengue, (2) trimester of which it developed had a significant association with adverse maternal outcomes (P < .001, P = .028). Presence of headache, nausea and vomiting showed an inverse relationship in the chi-squared test [Table 3]. Labour precipitation during or immediately following the dengue episode in the third trimester showed a significant association with dengue category (P = .013). Of continuous variables, (1) a lower platelet count and (2) higher AST level were significantly associated with adverse maternal outcomes (P<.001, P=.01) [Table 4].

**Table 02.**
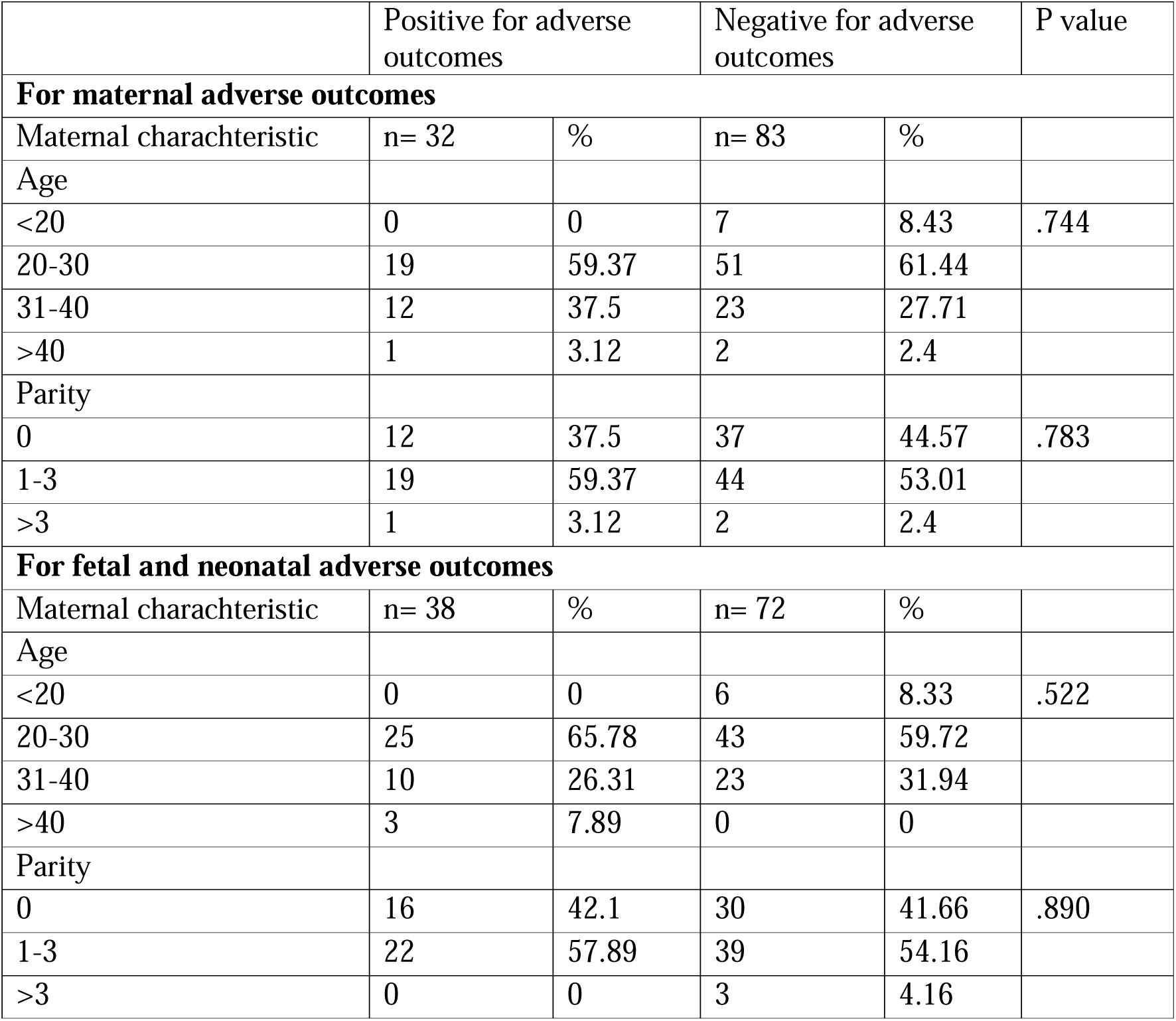
Distribution of confounders.

|  | Positive for adverse outcomes |  | Negative for adverse outcomes |  | P value |
| --- | --- | --- | --- | --- | --- |
| For maternal adverse outcomes |  |  |  |  |  |
| Maternal characteristic | n= 32 | % | n= 83 | % |  |
| Age |  |  |  |  |  |
| <20 | 0 | 0 | 7 | 8.43 | .744 |
| 20-30 | 19 | 59.37 | 51 | 61.44 |  |
| 31-40 | 12 | 37.5 | 23 | 27.71 |  |
| >40 | 1 | 3.12 | 2 | 2.4 |  |
| Parity |  |  |  |  |  |
| 0 | 12 | 37.5 | 37 | 44.57 | .783 |
| 1-3 | 19 | 59.37 | 44 | 53.01 |  |
| >3 | 1 | 3.12 | 2 | 2.4 |  |
| For fetal and neonatal adverse outcomes |  |  |  |  |  |
| Maternal characteristic | n= 38 | % | n= 72 | % |  |
| Age |  |  |  |  |  |
| <20 | 0 | 0 | 6 | 8.33 | .522 |
| 20-30 | 25 | 65.78 | 43 | 59.72 |  |
| 31-40 | 10 | 26.31 | 23 | 31.94 |  |
| >40 | 3 | 7.89 | 0 | 0 |  |
| Parity |  |  |  |  |  |
| 0 | 16 | 42.1 | 30 | 41.66 | .890 |
| 1-3 | 22 | 57.89 | 39 | 54.16 |  |
| >3 | 0 | 0 | 3 | 4.16 |  |

**Table 03.** Predictors of adverse outcomes – categorical variables.

|  | Maternal |  |  |  |  | Fetal and neonatal |  |  |  |  |
| --- | --- | --- | --- | --- | --- | --- | --- | --- | --- | --- |
|  | Positive for adverse outcomes |  | Negative for adverse outcomes |  | P value | Positive for adverse outcomes |  | Negative for adverse outcomes |  | P value |
| Predictor variable | n= 32 | % | n= 83 | % |  | n= 38 | % | n= 72 | % |  |
| Category of dengue |  |  |  |  |  |  |  |  |  |  |
| DF | 8 | 10.9 | 65 | 89 | < 0.001 | 20 | 28.9 | 49 | 71 | .194 |
| DHF | 21 | 53.8 | 18 | 46.1 |  | 16 | 42.1 | 22 | 57.8 |  |
| DES | 3 | 100 | 0 | 0 |  | 2 | 66.6 | 1 | 33.3 |  |
| Trimester |  |  |  |  |  |  |  |  |  |  |
| First trimester | 0 | 0 | 23 | 100 | .028 | 7 | 33.3 | 14 | 66.6 | .671 |
| Second trimester | 11 | 23.9 | 35 | 76 |  | 13 | 30.2 | 30 | 69.7 |  |
| Third trimester | 21 | 45.6 | 25 | 54.3 |  | 18 | 39.1 | 28 | 60.8 |  |
| Clinical |  |  |  |  |  |  |  |  |  |  |
| Days on admission > 2 | 8 | 30.7 | 18 | 69.2 | .703 | 9 | 37.5 | 15 | 62.5 | .730 |
| Days on admission ≤ 2 | 24 | 26.9 | 65 | 73 |  | 29 | 33.7 | 57 | 66.2 |  |
| Fever lasted > 4 days | 11 | 26.1 | 31 | 73.8 | .974 | 14 | 35 | 26 | 65 | .652 |
| Fever lasted ≤ 4 days | 18 | 26.4 | 50 | 73.5 |  | 20 | 30.7 | 45 | 69.2 |  |
| Highest T > 38 °C) | 23 | 25.8 | 66 | 74.1 | .379 | 28 | 32.5 | 58 | 67.4 | .406 |
| Highest T ≤ 38 °C) | 9 | 34.6 | 17 | 65.3 |  | 10 | 41.6 | 14 | 58.3 |  |
| Headache | 6 | 12.5 | 42 | 87.5 | .001 | 13 | 27.6 | 34 | 72.3 | .189 |
| Myalgia | 16 | 29.6 | 38 | 70.3 | .684 | 18 | 33.9 | 35 | 66 | .901 |
| Arthralgia | 15 | 27.2 | 40 | 72.7 | .899 | 18 | 33.9 | 35 | 66 | .901 |
| Respiratory symptoms | 7 | 38.8 | 11 | 61.1 | .254 | 6 | 33.3 | 12 | 66.6 | .905 |
| Retroorbital pain | 2 | 15.3 | 11 | 84.6 | .287 | 3 | 25 | 9 | 75 | .461 |
| Nausea and vomiting | 4 | 12.9 | 27 | 87 | .030 | 11 | 36.6 | 19 | 63.3 | .774 |
| Backpain | 9 | 33.3 | 18 | 66.6 | .465 | 10 | 38.4 | 16 | 61.5 | .630 |

(T – temperature, DF – dengue fever, DHF – dengue hemorrhagic fever, DES- dengue expanded syndrome)

**Table 04.** Predictors of adverse outcomes – continuous variables.

|  |  |  |  |
| --- | --- | --- | --- |
| Maternal |  |  |  |
| Predictor variable | Median - Positive for adverse outcomes | Median -Negative for adverse outcomes | p-value |
| Highest temperature | 38.5 | 38.7 | 1.0 |
| Lowest platelet count | 37.0 | 87.0 | <0.001 |
| Highest AST | 92.0 | 50.0 | 0.01 |
| Highest ALT | 52.0 | 34.0 | 0.34 |
| Fetal and neonatal |  |  |  |
| Highest temperature | 38.9 | 38.6 | 0.60 |
| Lowest platelet count | 65.5 | 69.5 | 0.53 |
| Highest AST | 69.5 | 57.0 | 0.20 |
| Highest ALT | 38.0 | 35.5 | 0.40 |

(AST- aspartate amino transferase, ALT-alanine amino transferase)

Owing to the small numbers, fetal and neonatal adverse outcomes were pooled together for the analysis of associations. None of the categorical or continuous variables showed a significant association with fetal or neonatal outcomes [Table 3,4].

## Discussion

The findings of this study shed light on the multifaceted challenges posed by antenatal dengue infection. The distribution of dengue infections throughout the year correlates with rainfall. This highlights the importance of implementing robust vector control measures to mitigate the spread of the disease.

Maternal adverse outcomes emerged as a significant concern, affecting over a quarter of the cases studied. These outcomes ranged from maternal death to serious complications such as dengue hepatitis and encephalitis, with ICU admission being the most common. This underscores the importance of not overlooking a possible dengue infection in a pregnant woman from an affected area or with a travel history to a tropical country. Multidisciplinary involvement is undeniable in cases of DES.

The maternal mortality ratio (1/101) in the study differed significantly from the general population ratio for the index year which is 39 per 100,000 live births (P <.001).^3^ In the case of the maternal death in this series, it is important to note the delay in presentation was a significant reason for the adverse outcome. This underscores the urgent need for prompt detection and intervention strategies to prevent fatalities associated with dengue infection during pregnancy.

A semi-ecological study in French Guiana associated dengue infection in the first trimester with post-partum hemorrhage (PPH) and preterm birth.^11^ However, none of the first-trimester episodes in our study were linked to maternal adverse outcomes, while precipitation of labour occurred in the third trimester, significantly associated with DHF compared to DF. Postpartum hemorrhage risk was low at 3.4%, with a 7.8% risk of needing a blood product transfusion, consistent with previous studies.^12, 13^

Factors associated with adverse maternal outcomes were dengue category, trimester of infection, and dropping of platelet count and rise of AST levels. This highlighs the potential utility of these parameters as early warning signs for healthcare providers managing antenatal dengue cases. Additionally, extracellular fluid loss, thrombocytopenia, and elevated liver enzymes could be indicative of pre-eclampsia or HELLP syndrome. Therefore, it is important to investigate a recent history of fever or travel history in cases with an atypical presentation of these common obstetric complications.

However, presence of headache, nausea and vomiting were negatively associated with maternal adverse outcomes. This may be due to the small sample size or retrospective nature of data collection. If a patient is severely ill on presentation, they may not have expressed these symptoms or may not have documented as the diagnosis was obvious.

Our study reported a significant difference in intrauterine fetal demise (IUD) rates compared to Sri Lankan national statistics, with similar findings to a prospective matched study.^3,14^ The risk of adverse fetal outcomes, including intrauterine demise (IUD) and miscarriages, highlights the potential impact of maternal dengue infection on fetal health and viability. Restrepo et al demonstrated a significant association between low birth weight and antenatal dengue infection in a retrospective cohort study.^15^ In contrast, risk of having a low birth weight (<2.5kg) was 12.8% (13/101) in the index study and it is not statistically different (P=.499) from the national value of 15.7% for the same year.^3^

Transplacental transmission of dengue infection was observed in a subset of cases, displaying the potential for vertical transmission and the need for close monitoring of neonates born to dengue-infected mothers. Mother-to-child transmission of dengue fever is well established in literature.^16,17^ While the percentage of cases with transplacental transmission was relatively low, the implications for neonatal health warrant further investigation, particularly regarding long-term sequelae and management strategies. Clinical or laboratory variables did not show promising predictability for fetal or neonatal adverse outcomes, possibly due to the time gap from infection to delivery and small numbers precluding subgroup analysis. This highlights the complexity of dengue infection during pregnancy and emphasizes the need for further research to exemplify the underlying mechanisms and risk factors associated with adverse fetal and neonatal outcomes.

Key limitations of this study include its retrospective design and lack of an unexposed comparator group leaving a potential for residual confounding. The lack of consistent documentation regarding the method of gestational age estimation further complicates the interpretation of findings related to trimester-specific outcomes. Additionally, the small sample size may limit the generalizability of the results, warranting further investigation in larger, prospective studies.

In conclusion, antenatal dengue infection poses significant risks to maternal, fetal, and neonatal health. However, further research is needed to elucidate the complex interplay of factors influencing fetal and neonatal outcomes and to develop targeted management strategies to optimize pregnancy outcomes in the context of dengue infection.

## Data Availability

All data produced in the present study are available upon reasonable request to the authors

## Reference

1. World Health Organization [homepage on the internet]. Dengue guidelines for diagnosis, treatment, prevention and control: new edition; 2009. [cited 2024 Feb 20] Available from: https://apps.who.int/iris/handle/10665/44188

2. Sri Lanka Epidemiology Unit [homepage on the internet]. Trends; 2020. [cited 2024 Feb 20] Available from: https://www.epid.gov.lk/web/index.php?option=com_casesanddeaths&section=trends&Itemid=448&lang=en

3. Family Health Bureau [homepage on the internet]. Annual Report; 2017. [cited 2024 Feb 20] Available from: https://fhb.health.gov.lk/annual-report/

4. World Health Organization [homepage on the internet]. Dengue fever – Sri Lanka; 2017. [cited 2024 Feb 20] Available from: https://www.who.int/emergencies/disease-outbreak-news/item/19-july-2017-dengue-sri-lanka-en

5. Sri Lanka Epidemiology Unit [internet]. Guidelines on management of DF/DHF in adults; 2012. [cited 2024 Feb 20] Available from: https://www.dengue.health.gov.lk/web/images/Guidelines_on_management_of_dengue_fever_and_dengue_haemorrhagic_fever_in_adults_1.pdf

6. Sri Lanka Epidemiology Unit [homepage on the internet]. Guidelines on management of DF/DHF in adults; 2012. [cited 2024 Feb 20] Available from: https://www.dengue.health.gov.lk/web/images/Guidelines_on_management_of_dengue_fever_and_dengue_haemorrhagic_fever_in_adults_1.pdf

7. Pipkin FB. Maternal physiology. In: Edmonds DK, Lees C, Bourne T, editors. Dewhurst’s text book of Obstetrics and Gynaecology. New Jersey: Wiley-Blackwell; 2018. p.1–17

8. Jameson L, Fauci AS, Kasper DL, Hauser SL, Longo DL, Loscalzo J. Harrison’s Principles of Internal Medicine. 20th ed. New York: McGraw-Hill; 2018.

9. American College of Obstetricians and Gynecologists [internet]. How Your Fetus Grows During Pregnancy; 2024. [cited 2024 Feb 20] Available from: https://www.acog.org/womens-health/faqs/how-your-fetus-grows-during-pregnancy

10. Nelson-Piercy C. Handbook of Obstetric Medicine. 6th ed. Florida: CRC Press; 2001.

11. Hanf M, Friedman E, Basurko C, Roger A, Bruncher P, Dussart P et al. Dengue epidemics and adverse obstetrical outcomes in French Guiana: a semi-ecological study. Trop Med Int Health. 2014;19:153–8.

12. Carles G, Talarmin A, Peneau C, Bertsch M. Dengue fever and pregnancy: A study of 38 cases in french Guiana. J Gynecol Obstet Biol Reprod (Paris) 2000;29:758–2.

13. Basurko C, Carles G, Youssef M, Guindi WEL. Maternal and fetal consequences of dengue fever during pregnancy. Eur J Obstet Gynecol Reprod Biol 2009;147:29–2.

14. Basurko C, Everhard S, Matheus S, Restrepo M, Hildéral H, Lambert V. et at. A prospective matched study on symptomatic dengue in pregnancy. PLoS ONE 2018;13(10): e0202005.

15. Restrepo BN, Isaza DM, Salazar CL, Ramírez JL, Upegui GE, Ospina M. Prenatal and postnatal effects of dengue infection during pregnancy. Biomed Rev Inst Nac Salud 2003;23:416–3.

16. Tan PC, Rajasingam G, Devi S, Omar SZ. Dengue infection in pregnancy: prevalence, vertical transmission, and pregnancy outcome. Obstet Gynecol 2008;111:1111–7.

17. Phongsamart W, Yoksan S, Vanaprapa N, Chokephaibulkit K. Dengue virus infection in late pregnancy and transmission to the infants. Pediatr Infect Dis J 2008;27:500–4.

